# Protocol for Project Recovery: Cardiac Surgery - Leveraging Digital Platform for Efficient Collection of Longitudinal Patient-Reported Outcome Data Towards Improving Postoperative Recovery

**DOI:** 10.1101/2020.01.12.20017269

**Authors:** Makoto Mori, Cornell Brooks, Erica Spatz, Bobak J Mortazavi, Sanket S. Dhruva, George C. Linderman, Lawrence A. Grab, Yawei Zhang, Arnar Geirsson, Sarwat I. Chaudhry, Harlan M. Krumholz

## Abstract

**Introduction:** Improving postoperative patient recovery after cardiac surgery is a priority, but our current understanding of individual variations in recovery and factors associated with poor recovery is limited. We are using a health-information exchange platform to collect patient-reported outcome measures (PROMs) and wearable device data to phenotype recovery patterns in the 30-day period after cardiac surgery hospital discharge, to identify factors associated with these phenotypes and to investigate phenotype associations with clinical outcomes.

**Methods and analysis:** We designed a prospective cohort study to enroll 200 patients undergoing valve, coronary artery bypass graft, or aortic surgery at a tertiary center in the U.S. We are enrolling patients postoperatively after the intensive care unit (ICU) discharge, and delivering electronic surveys directly to patients every 3 days for 30 days after hospital discharge. We will conduct medical record reviews to collect patient demographics, comorbidity, operative details and hospital course using the Society of Thoracic Surgeons (STS) data definitions. We will use phone interview and medical record review data for adjudication of survival, readmission, and complications. We will apply group-based trajectory modeling to the time-series PROM and device data to classify patients into distinct categories of recovery trajectories. We will evaluate whether certain recovery pattern predicts death or hospital readmissions, as well as whether clinical factors predict a patient having poor recovery trajectories. We will evaluate whether early recovery patterns predict the overall trajectory at the patient-level.

**Ethics and dissemination:** The Yale Institutional Review Board approved this study. Following the description of the study procedure, we obtain written informed consent from all study participants. The consent form states that all personal information, survey response, and any medical records are confidential, will not be shared, and are stored in an encrypted database.

**Strengths and limitations of this study:**

- This study will assess the patient perspective on recovery after cardiac surgery at a high frequency within the 30-day postoperative period with surveys and activity monitoring via a health information platform and wearable devices.
- Using longitudinal patient-reported outcomes measure (PROM) data, this study will define recovery patterns and factors associated with different recovery trajectories and guide the development interventions to improve recovery and support expansion of the study to additional sites.
- The study is single center and the sample size is limited.

## Background

Improving postoperative patient recovery is a priority. Readmission rates in the post-operative period are high. Moreover, in the United States, the expansion of episode-based payments and performance measures is increasing interest in the post-acute experience of patients^1, 2^. However, we generally lack systematically-collected information on the experience of patients in the post-acute period, as few studies rigorously collecting information using established patient-reported outcomes measures (PROMs). We have, for example, little information about the variation of the trajectories of recovery and the factors most strongly associated with better outcomes^3^.

The assessment of the patient experience can provide important insights into the process of recovery that is not evident through clinical outcomes or intermittent clinical office visits. PROMs and wearable devices can provide complementary information by providing measurements of how the patient’s experience and functional status change over time^4^. Current digital platforms allow us to efficiently collect PROMs and wearable-generated data at high frequencies and with little cost and burden. These automated data collection approaches may minimize the bias introduced by clinician-directed patient interviews^5^. Such a platform is highly suited to obtain repeated measures to characterize a time-dependent process such as recovery^6^.

Cardiac surgery is an ideal area for the study of recovery. Many patients have good outcomes, but the limited existing evidence suggests a wide variation in the post-operative experience of these patients^7^. However, these patients’ experience has been poorly studied, as most studies of recovery simply assess deaths and complications.

Characterizing the recovery from the patient perspective is important for many reasons. First, shared decision-making and informed consent should be guided not only by the risk of mortality and complications but also by the recovery experience. Understanding variations in recovery could enable the early identification of people who are struggling and require additional attention. Recovery data from the patient perspective may enable remote monitoring after the procedure to selectively and preemptively intervene on those at high risk of poor recovery to improve outcomes. Characterization of recovery can also be used to identify patient, surgeon, procedural, and institutional factors that are associated with different patterns. With this information we can identify modifiable risk factors for poor recovery.

Thus, at this juncture, there are several notable gaps in knowledge. First, although recovery occurs over time, most studies of recovery included a small number of timepoints, and the recovery trajectory phenotypes remains poorly defined^3^. Cohort-level average of recovery trajectories is a common way of reporting^3^ and can indicate how patients recover on average^7^, but it obscures individual variation such as rapid early recovery, gradual recovery, or initial recovery followed by a decline. Second, we have limited understanding of how recovery trajectories vary by patient factors, operation types, center or surgeon characteristics, procedural processes, and complications, which limit opportunities to identify high risk patients preemptively and intervene.

Accordingly, our overall objective is to characterize short-term trajectories of patient recovery after cardiac surgery using PROMs and wearable data. We are conducting a prospective study to characterize trajectories of postoperative recovery in multiple domains after cardiac surgery. The specific aims of this study are to: 1) leverage a digital data platform to collect PROM and wearable device data to bring forth the variable individual recovery trajectories, 2) describe distinct classes of recovery trajectories and clinical factors associated with the classes, and 3) to evaluate whether early postoperative recovery trajectory predicts later recovery trajectory. In addition, we will investigate optimal ways to manage missing data specific to these time-series data This study is a step toward using this approach to prospectively monitor and preemptively identify patients at risk of poor recovery and facilitate intervention to reduce the risk of adverse events.

## Methods

### Design Overview

This is a prospective cohort study of patients who are undergoing valve, CABG, or aortic surgery at a tertiary center in the U.S. We chose the operations because they are the most common cardiac operations performed^8^. We are enrolling patients postoperatively after ICU discharge in order to ensure clinical stability, and we electronically delivering surveys directly to patients every 3 days for 30 days after hospital discharge to study patient trajectories in multiple domains characterizing recovery. The closing phone interview after 30 days, electronic medical record review, and linkage to the Society of Thoracic Surgeons database are used to confirm survival, readmission, and complications. The closing interview asks about details of readmissions if they occurred, patients’ overall satisfaction with the study, and whether their experience was well captured by the summary of their PROM data. We will apply group-based trajectory modeling to the longitudinal PROM data to identify distinct categories of recovery trajectories in a data-driven fashion. We also identify predictors of protracted recovery trajectory and evaluate whether early recovery patterns (<10 days) predict the overall trajectory (30 days) at the patient-level. The Yale Institutional Review Board approved this study.

### Patient Population

This study began in January 2019 and is ongoing. The study is taking place at Yale-New Haven Hospital, a tertiary center in the United States, where over 1,100 cardiac surgeries are performed annually. Inclusion criteria are patients of age 18 and older who are undergoing coronary artery bypass grafting (CABG), valve replacement or repair, or aortic operations. Exclusion criteria are those who undergo heart transplant, extracorporeal membrane oxygenation (ECMO), adult congenital operations, or ventricular assist device implantation, as these patient populations tend to have a longer course of intensive care unit stay^9^, precluding the timely enrollment necessary to capture immediate postoperative recovery. We also excluded those who do not own a smartphone or a tablet or those who do not speak or read English, because the digital platform for PROM data collection relies on patients responding to surveys displayed on web browser via email or text, and the surveys were written in English language. We do not allow proxy for survey response and consequently excluded patients who were not able to respond by themselves as determined by the research assistant.

### Recruitment

Recruitment takes place postoperatively after the patient has left the intensive care unit (ICU) for the step-down or floor unit (Figure 1). We chose to enroll patients postoperatively, as opposed to preoperatively, because postoperative enrollment allows for enrollment of patients who undergo surgery under non-elective settings. Recruitment after transfer from the ICU setting ensures clinical stability. A research assistant (RA) visits the patient and after confirming the patient is eligible to participate and following the description of the study procedure, obtains written informed consent (Supplementary Material S1) from all study participants. The informed consent form states that all personal information, survey response, and any medical records are confidential, will not be shared, and will be stored in an encrypted database.

**Figure 1:**
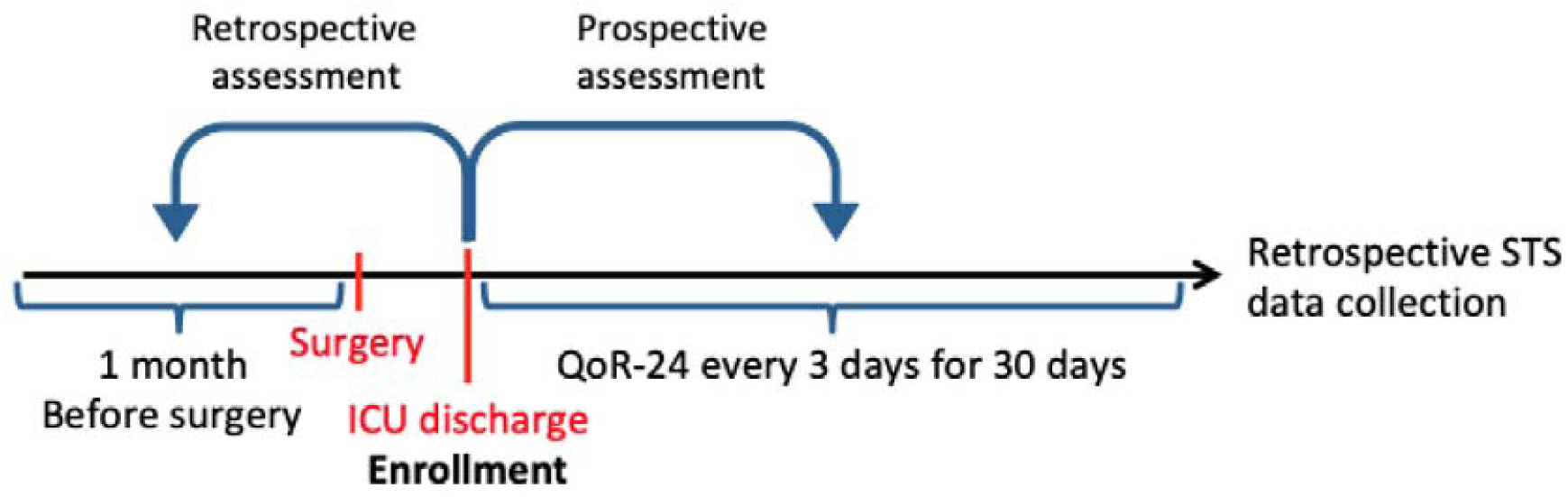
Timing of patient enrollment and PROM administration. The figure shows the timing of patient enrollment and PROM administration over the clinical course. Baseline function is assessed by retrospectively asking the patient about their state of health during 1 month prior to the operation. 24-item Quality of Recovery questionnaire is administered every 3 days for 30 days following discharge from the intensive care unit.

We iteratively refined the enrollment process to minimize the onboarding time, which includes obtaining informed consent and signup process directed by the RA on a tablet device to enter patient name and email address or phone number and takes approximately 10-15 minutes.

### PROM instrument and administration

We use 24-item quality of recovery (QoR-24) to characterize patients’ postoperative recovery in various domains. The questionnaire consists of 24 items that were developed and validated in inpatient and outpatient surgical populations^10-13^. The instrument was previously adapted into a mobile format and was successfully used to administer the survey daily for 14 days^11, 12^. We added 3 items to QoR-24 to capture the self-reported time patients went to sleep, the time they awakened, and their global perception of how much they have ‘recovered’ in a 0-100% scale. The resulting 27-item questionnaire takes 2-4 minutes to complete, making its frequent administration feasible (Supplementary Material S2). Among the published studies in cardiac surgery, this study will have the highest number of PROM data points collected in the first postoperative month^3^.

### Digital data platform

We are delivering surveys on the day of enrollment and every 3 days for 30 days. This method provides detailed longitudinal data across multiple domains of recovery (Figure 2). To facilitate data organization and scheduled survey delivery, we use Hugo (Me2Health, LLC, Guilford CT, USA) a patient-centered health data sharing platform, which has a customizable survey delivery function and reminder feature to facilitate data collection. Hugo platform allows for automated delivery of surveys without researchers having to directly contact patients, which facilitates high-frequency data collection. Additionally, it imports data from connected wearable devices to facilitate centralization of patient health data. The patients retain access to their own data in a cloud-based account.

**Figure 2:**
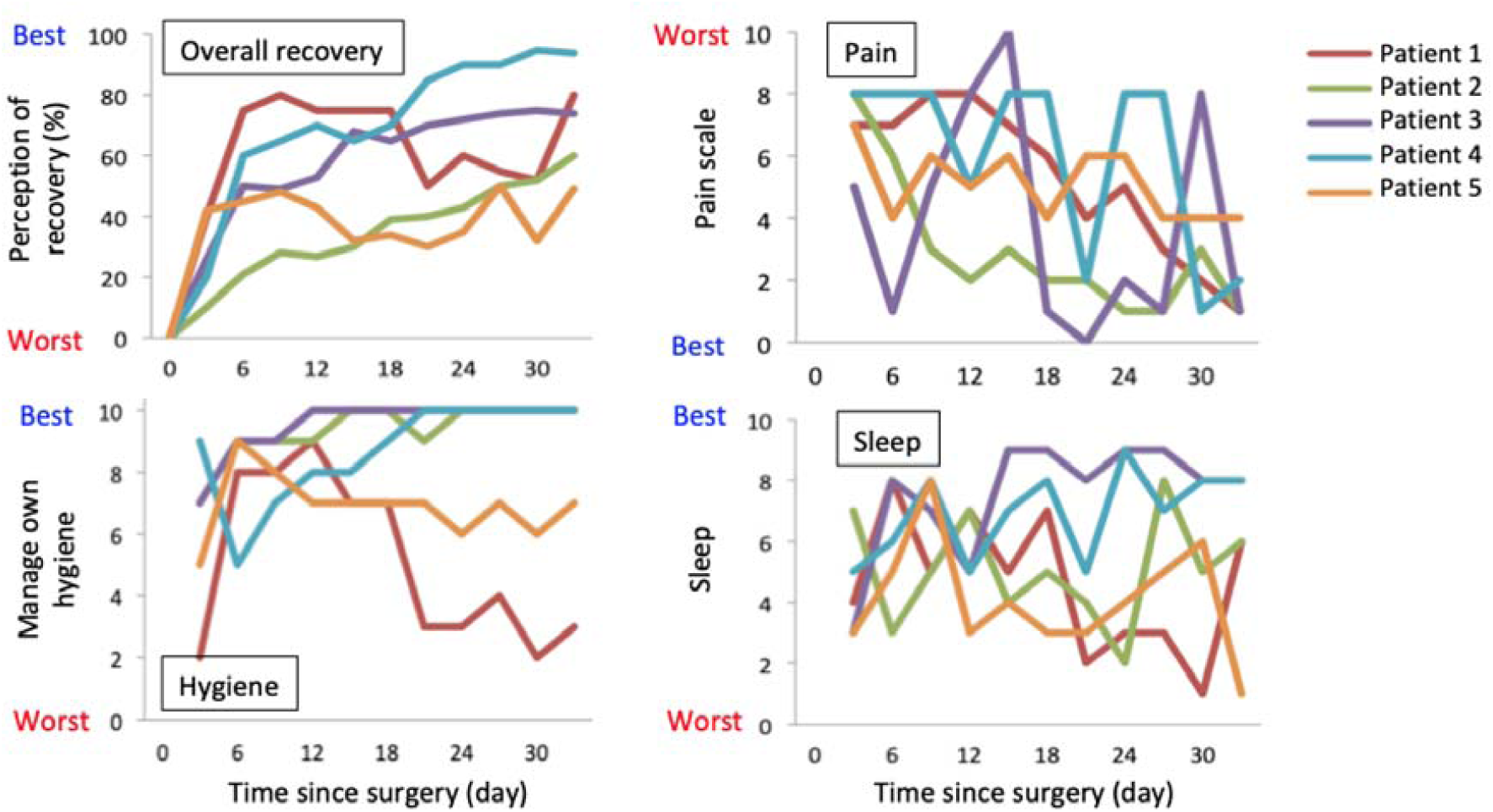
Sample trajectories of recovery in 5 patients. The figures display trajectories of recovery in different domains in 5 patients. Each color corresponds to the same patient. Overall recovery is the patient’s perception of overall recovery in 0 to 100% scale. Pain in surgical site is reported in 0 to 10 point scale, with 10 representing the worst pain. Being able to take care of own hygiene is reported in 0 to 10 point scale, with 10 representing complete independence in managing own hygiene. Patient’s perception of sleep quality is reported in 0 to 10 point scale, with 10 being the best sleep.

### Identifying common reasons for low response rate

Recognizing that the survey response will be incomplete for some participants, we have conducted a phone interview with the first 22 patients to learn reasons for low responses and identify strategies to minimize the barriers toward survey response for subsequent participants. In the first 22 patients, we identified 5 with response rate of <50% and conducted recorded phone interviews. Our interview guide (Supplementary Material S3) contained questions to elucidate technical barriers, differential preferences for engagement, and or any other issues precluding survey completion. We also asked whether the length of the questionnaire or types of questions asked made it difficult to complete the survey. Two members of the research team (CB and MM) evaluated the interview recordings to identify common reasons for low response rate. This suggested the potential importance of reminder to maintain patient engagement. We modified the protocol to contact all participants approximately 10 days after enrollment.

### Additional clinical data and adjudication of hospitalization and survival

Additionally, we are using the Society of Thoracic Surgeons (STS) Adult Cardiac Surgery Database data specifications to retrospectively collect clinically relevant data in this patient population. Pre-specified candidate predictors in this database will be used to identify clinical predictors of recovery trajectories (Table 1). The STS database contains patient demographics, comorbidities, presenting clinical status, operative details, and postoperative mortality and morbidity up to 30 days after the time of operation^14^. These data are routinely collected at Yale New Haven Hospital.

**Table 1:**
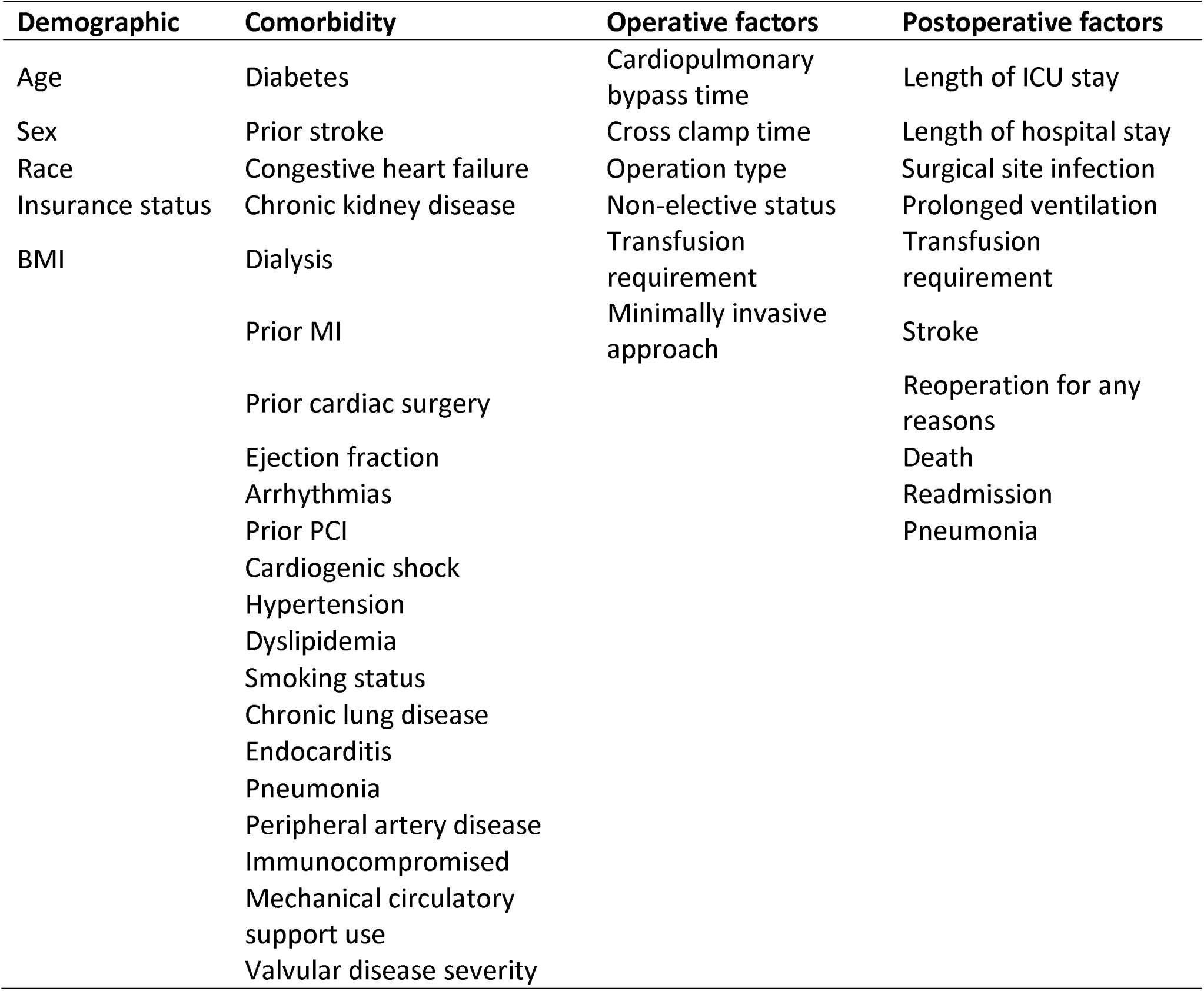
Candidate predictors of recovery trajectory

| Demographic | Comorbidity | Operative factors | Postoperative factors |
| --- | --- | --- | --- |
| Age | Diabetes | Cardiopulmonary bypass time | Length of ICU stay |
| Sex | Prior stroke | Cross clamp time | Length of hospital stay |
| Race | Congestive heart failure | Operation type | Surgical site infection |
| Insurance status | Chronic kidney disease | Non-elective status | Prolonged ventilation |
| BMI | Dialysis | Transfusion requirement | Transfusion requirement |
|  | Prior MI | Minimally invasive approach | Stroke |
|  | Prior cardiac surgery |  | Reoperation for any reasons |
|  | Ejection fraction |  | Death |
|  | Arrhythmias |  | Readmission |
|  | Prior PCI |  | Pneumonia |
|  | Cardiogenic shock |  |  |
|  | Hypertension |  |  |
|  | Dyslipidemia |  |  |
|  | Smoking status |  |  |
|  | Chronic lung disease |  |  |
|  | Endocarditis |  |  |
|  | Pneumonia |  |  |
|  | Peripheral artery disease |  |  |
|  | Immunocompromised |  |  |
|  | Mechanical circulatory support use |  |  |
|  | Valvular disease severity |  |  |

We will determine mortality and hospital readmissions by several approaches: review of hospital records, review of cardiac surgery clinic notes, and conducting closing phone interviews with the patient or contact person previously identified.

### Patient Involvement

Prior to launching the study, we interviewed 5 patients both in pre and postoperative settings to evaluate whether the frequency of survey delivery and PROM instrument were likely to adequately capture their experience of recovery. All patients agreed that the frequency of questionnaire administration and the length of the PROM instrument were reasonable and provided face validity that the questionnaire captured aspects of recovery that were important to the patients. Additionally, this article is authored with a patient (LG) who participated in the study to reflect his perspective on the study design and experience in responding to the surveys.

### Sample size

The study sample target is 200 patients. Adequate sample size for studies using group-based trajectory modeling depends on the dataset’s representativeness of the population of interest^15^. Therefore, the concept of statistical power traditionally used for sample size calculation does not apply to latent class analyses. We may generate a larger simulation dataset from the measured patient trajectory data to perform a split-sample testing, evaluating whether trajectories generated from the derivation sample would allow for satisfactory categorization of the testing dataset. Additionally, the study setting is scalable to increase the sample size by increasing the enrollment period, should a larger sample size become necessary.

### Analytical approach – group-based trajectory modeling

The resulting dataset is a complex time-series data, with each patient having 10 data points (one every three days) at different postoperative times for each item. A practical approach to dimension reduction is group-based trajectory modeling, which is a type of latent class analysis that groups similar patient trajectories according to a number of features derived from the time-series data^16, 17^. This approach allows for dimension reduction of the complex time-series data into several distinct classes of recovery trajectories. These trajectories can be labeled according to the observed clinical phenotype of trajectories, for example ‘fast recovery,’ ‘average recovery,’ or ‘protracted recovery,’. This data-driven categorization enables additional regression modeling to identify predictors of patients belonging to a certain class of recovery path.

The dataset will be classified into distinct categories of trajectories at domain level, using group-based trajectory modeling^16, 17^. Traj package on R^18^ or Proc Traj package on SAS^15^, performs trajectory modeling by first extracting 24 features of patient-level trajectory, selecting a subset of features that describes the overall trajectory, and identifying optimal number of classes to group the trajectories based on the longitudinal k-means method. The 24 features include range, mean change per unit time, and slope of the linear model (Table 2), which have been demonstrated to discriminate between stable-unstable, increasing-decreasing, linear-nonlinear, and monotonic-nonmonotonic patterns of trajectories^18^. K-means method partitions the time-series data into k groups such that the mean squared error distance of each data point from the assigned cluster is minimized^19^. The optimal number of clusters is determined by the minimization of Bayesian information criterion, which signifies the balance between model’s complexity and the ability to describe the dataset. This process yields distinct classes of patient trajectories in a data-driven fashion. Trajectories will be identified separately for the 5 domains and 1 global recovery measure.

**Table 2:**
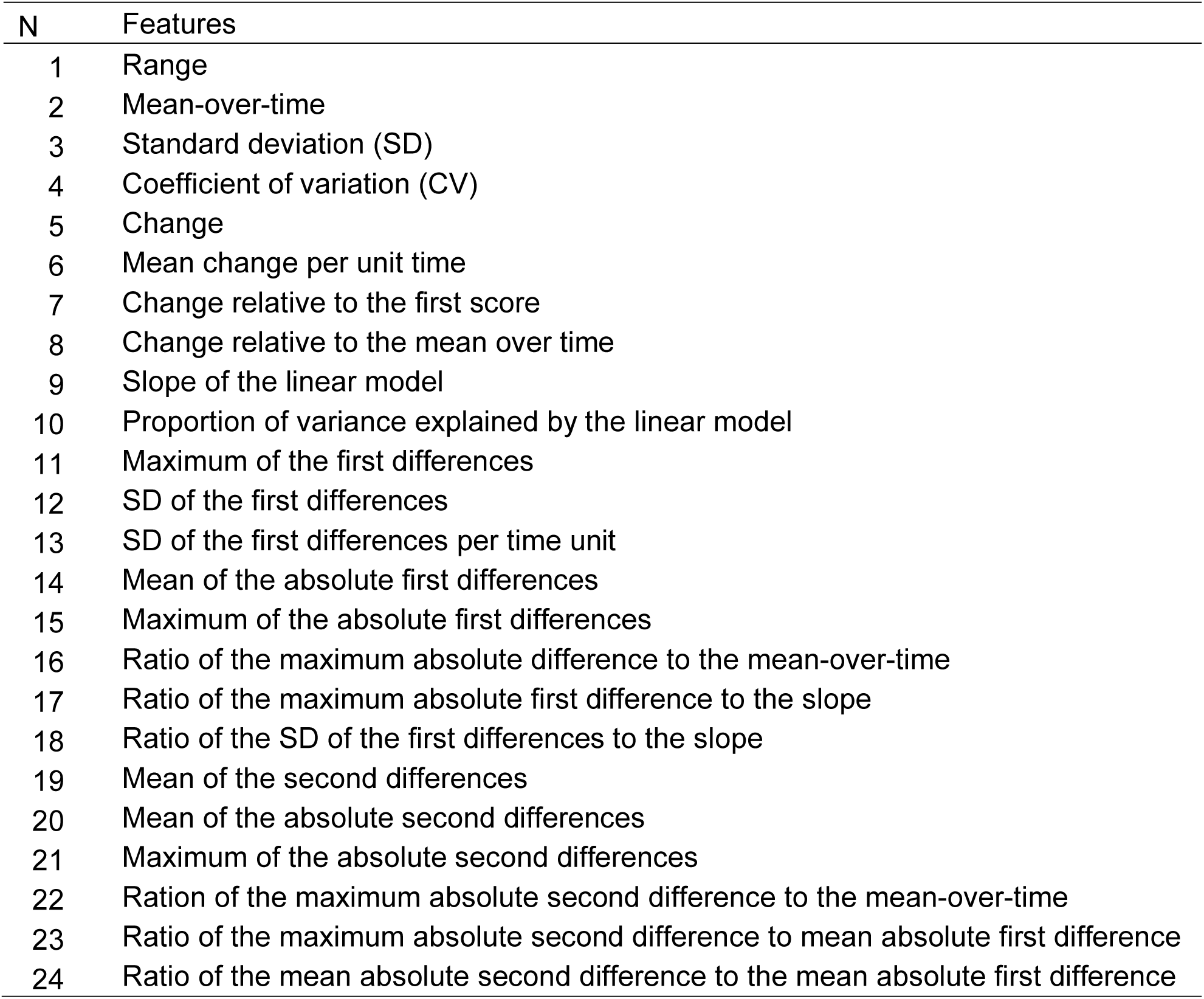
24 features of trajectory used in group-based trajectory model

### Analytical approach – missing data

Because missing data are inevitable in longitudinal PROMs, there is a need employ an appropriate handling of missing data. Multiple imputation prior to latent class analysis may yield a less biased estimate of the resulting trajectories. An alternative approach used in group-based trajectory models assumes the data are missing at random (MAR) and generates the maximum likelihood of the model parameters^20^. MAR is valid when the response attrition is independent of the group membership. However, patient attrition is oftentimes dependent on clinical characteristics and likely related to the class of trajectory itself. An extension of the model allows for modeling of attrition across trajectory groups^21^, permitting dropout probability to vary as a function of covariates or observed outcomes prior to dropout and yields a more robust estimate of the probability of group membership. As such, we will perform sensitivity analysis to compare the trajectories generated via raw data vs. data preprocessed with multiple imputation vs. trajectories generated via trajectory model accounting for response attrition.

## Results

Between January and May 2019, we have enrolled 22 patients who completed the 30-day follow-up. In this cohort, median age was 58.5 years (interquartile range 53.5-67.0) and 7 (32%) were women. There were 9 (41%) mitral valve repair cases and 6 isolated or concomitant CABG (27%).

### Barriers to completing surveys

Of the 22 patients enrolled, 3 (14%) did not complete any surveys, 19 (86%) completed at least 3 surveys, and 17 patients (77%) completed at least 6 of 11 delivered surveys (>50% of delivered surveys). Of the 5 patients who completed less than half of the surveys, we successfully contacted 4, and 1 could not be reached after 5 attempts. All 4 reported that the major barriers precluding survey completion were their clinical conditions: 2 described readmissions as an overwhelming event that made them feel continuing survey participation challenging, and 2 described not feeling well in general, which precluded participation. All 4 patients noted that text or email reminders might have been helpful to sustain participation. Based on these responses, we modified the protocol to contact all participants approximately 10 days after enrollment to improve engagement and resolve any patient-specific issues in completing the surveys.

### Clinical outcomes

There were no deaths during follow-up. Two (9%) patients experienced at least 1 hospital readmission. Figure 2 depicts the breadth in recovery trajectories in pain, sleep, ability to take care of own hygiene, and perception of overall recovery in five patients with complete response.

### Patient perspective

An author (LG) participated in this study as he underwent cardiac surgery. He noted that the length and frequency of the questionnaire was reasonable and helped him to be more aware of the recovery process because responding to the questions facilitated introspection on his progress across different domains. He recommended that the study platform feedback the data to study participants, such as a visual summary of the trajectory or response for patients to better gauge their progress. Additionally, he noted that responses may differ across the time of the day, as he was able to better function physically in the afternoon compared with in the morning. Finally, he suggested the potential value of reviewing the questionnaire during preoperative counseling to highlight important aspects of recovery to provide better patient and family expectations. Investigators plan on providing a brochure to the clinic that contains this information.

## Discussion

This study will provide time-series data on short-term recovery after cardiac surgery using PROM instruments complemented by clinical records obtained via the STS database and electronic health records. This study will provide one of the highest density of postoperative PROM data in existing cardiac surgery literature^3^, and it will characterize the variability in individual recovery processes with a high temporal resolution. This study will be important in closing knowledge gaps around patient-level variations in trajectories because prior studies have mostly focused on changes in PROM scores at a limited number of time points^3^ or reporting group-level aggregate of longitudinal recovery data^7, 22^. Because recovery is an individual, variable, and time-dependent process, we designed our data collection and analytical approach to capture such features important to recovery.

This study has the potential to make a variety of contributions toward improving post-acute phase of care. First, we will be able to develop a preliminary nomogram of postoperative recovery for each domain and overall perception of recovery, which would be instrumental for patients and clinicians to gauge the breadth of possible recovery trajectories to facilitate informed shared decision-making. Second, identifying predictors of accelerated or protracted recovery, as classified by group-based trajectory model, may allow for individualized prediction of the postoperative recovery course to better inform the patients and family members. Third, early detection of recovery signals related to adverse events, such as mortality and readmission, may eventually facilitate preemptive intervention and focused monitoring of patients at an elevated risk for such events. Our design of the longitudinal PROM data collection allows for incremental update of such prediction as patients progress through the phase of recovery.

There are many challenges to the successful acquisition of patient measurements during recovery: efficient administration of PROMs in a way that does not require prohibitive amount of resources, minimizing selection bias originating from barriers to survey completion, handling of missing data that inevitably occurs in PROMs, and summarizing the complex data in a way that is interpretable to surgeons and patients^23^. Additionally, the use of wearables and device data require active patient participation in periodically charging the device, wearing them correctly, and reliably syncing the device to the server for data uploads. Moreover, there is a need to provide value to the patients for providing their recovery profile, such as giving them access to their health data in a meaningful way.

The resulting data collection, analytical, and output platforms have the potential of being implemented in the clinical setting where an integration of incrementally increasing PROM and clinical data provides the near-real time estimate of individual patient risk of adverse post-operative events. Such a model may allow for triggering of preemptive clinical intervention. An output may assimilate a form of clinical dashboard within the electronic health record system, which may be monitored at a centralized location where a trained clinician reviews high-risk cases filtered by the algorithm to further evaluate whether the patient condition warrants an intervention. Together, this workflow has a tremendous potential to improve post-acute phase of care following surgery.

### Lessons Learned from the initial experience

Through this first group of enrolled patients, we learned that most of the patients approached were willing to participate and consented to the study. By streamlining the enrollment process, the enrollment time shortened from over 1 hour on the first patient to approximately 10-15 minutes for the current enrollment. The overall response rate is acceptable, with 77% of the participants completing more than half of the delivered surveys independently without any intervention by researchers. Challenging recovery course, including readmissions may have interfered with patient engagement. While this would have resulted in an underrepresentation of those with protracted recovery or with complications, our preliminary data show we were able to capture variations in the trajectories of recovery.

To sustain patient engagement through challenging recovery course, we implemented a protocol for a research assistant to call the patient around 10 days after enrollment to troubleshoot any issues and reemphasize the importance of their participation. We believe that once the survey becomes part of clinical workflow with clinicians monitoring and responding to the PROM response, patient response rate would improve further.

We modified the enrollment protocol to reduce the enrollment time, because to some patients, the complexity and prolonged time spent for enrollment discouraged signups. Initial protocol for enrollment required patients to download an app and register. This resulted in a wide range of time spent for enrollment between 15 minutes and 90 minutes, with longer enrollment owing to technical challenges. These challenges include patients forgetting the password for app download, having to reset the password, and not having immediate email access to check account confirmation emails. Because our cardiac surgery patient population tended to be older, these technical challenges may have been pronounced. By not including the app download and allowing for the research assistant to enroll the patient via an online form with their permission, the enrollment time shortened significantly to 10-15 minutes.

Examining the initial individual data on recovery, there were wide variations in the trajectories of recovery even among only 5 patients. The variation suggests that the instrument we used was sensitive to capturing such differences. We also noted variations in improvement over time across different domains of recovery, where overall perception of recovery seemed to have a steady improvement pattern, while pain varied between consecutive measurements in some patients.

### Limitations

There are several limitations to this study. First, the single-center tertiary care setting limits the sample size and applicability of the findings to patients cared for in different settings. A multi-center study following the current study would address this limitation and evaluate whether the findings at our center are comparable to findings in other centers. Additionally, group-based trajectory modeling will classify patients into distinct trajectories based on similar recovery patterns, and this analytical approach may allow for generalization of the variations in the trajectories as long as our sample represents the breadth of the possible variation in recovery.

Another limitation is the exclusion of patients who cannot participate for various reasons. The use of digital platform is advantageous in reducing the resource intensity for data collection, but leads to exclusion of patients who do not own mobile devices, which likely affects older patients disproportionately. As the number of adults using mobile devices is increasing^24^, we believe this will become less of a limitation over time. Initiating this study now despite this limitation is important to establish a platform that may become the standard of postoperative care when the vast majority of patient population own digital devices in a predictably near future. Those who cannot participate due to lack of interest represent an important population that may be distinct in characteristics and risk profiles. We plan on minimizing the non-participation for the lack of interest by intermittent phone check-ins to sustain interests and identify barriers to inform strategies to increase engagement. In following studies, we may consider other forms of incentives to participate, if this population is indeed distinct and large in proportion. Additionally, when the PROM data are integrated into routine clinical care, patient engagement will likely increase substantially because they will be more inspired to share these data if they are used by their clinicians.

Finally, postoperative enrollment and retrospective assessment of preoperative health status, as opposed to preoperative enrollment, may introduce recall bias. We decided on postoperative enrollment, because preoperative enrollment precluded standardized enrollment of patients operated on under non-elective settings. Given the retrospective assessment of baseline health status takes place on the first postoperative survey, we believe the recall bias is minimized owing to the temporal proximity.

## Conclusion

This study will generate highly granular, longitudinal PROM data to characterize individual trajectories of patient recovery after cardiac surgery. Digital data sharing platforms promise to minimize the patient and researcher burden in administering and completing PROMs, allowing for characterization of granular progression of patients’ state of health over time in the postoperative period. Implementation of such study is complex but feasible, and it will serve as an important platform to facilitate clinical use of PROM data to improve the overall patient recovery.

## Data Availability

This is a study protocol article and the data are not available.

## Authors’ contributions

MM, HMK, SD, and AG developed the study and research question. MM and HMK developed analytical strategy with inputs from BJM, GCL, and YZ. SIC and ES guided refining the enrollment strategy and interpretation of the phone interview responses. LAG provided patient perspective on the study protocol and interpretation of the preliminary results. All authors developed and approved the final manuscript before submission.

## References

1. Wadhera RK, Yeh RW and Joynt Maddox KE. The Rise and Fall of Mandatory Cardiac Bundled Payments. JAMA. 2018;319:335–336.

2. Khera R, Dharmarajan K, Wang Y, Lin Z, Bernheim SM, Normand ST and Krumholz Hm. Association of the Hospital Readmissions Reduction Program With Mortality During and After Hospitalization for Acute Myocardial Infarction, Heart Failure, and Pneumonia. JAMA Netw Open. 2018;1:e182777.

3. Mori M, Angraal S, Chaudhry SI, Suter LG, Geirsson A, Wallach JD and Krumholz HM. Characterizing Patient-Centered Postoperative Recovery After Adult Cardiac Surgery: A Systematic Review. J Am Heart Assoc. 2019;8:e013546.

4. Gill TM, Gahbauer EA, Han L and Allore HG. Trajectories of disability in the last year of life. N Engl J Med. 2010;362:1173–80.

5. Pakhomov SV, Jacobsen SJ, Chute CG and Roger VL. Agreement between patient-reported symptoms and their documentation in the medical record. Am J Manag Care. 2008;14:530–9.

6. Moore FD. Getting well: the biology of surgical convalescence. Ann N Y Acad Sci. 1958;73:387–400.

7. Diab MS, Bilkhu R, Soppa G, Edsell M, Fletcher N, Heiberg J, Royse C and Jahangiri M. The influence of prolonged intensive care stay on quality of life, recovery, and clinical outcomes following cardiac surgery: A prospective cohort study. J Thorac Cardiovasc Surg. 2018;156:1906–1915.e3.

8. Thourani VH, Badhwar V, Shahian DM, O’Brien S, Kitahara H, Vemulapalli S, Brennan JM, Habib RH, Fernandez F, D’Agostino RS, Lobdell K, Rankin JS, Gammie JS, Higgins R, Sabik J, Schwann TA and Jacobs JP. The Society of Thoracic Surgeons Adult Cardiac Surgery Database: 2019 Update on Research. Ann Thorac Surg. 2019;108:334–342.

9. Blanche C, Blanche DA, Kearney B, Sandhu M, Czer LS, Kamlot A, Hickey A and Trento A. Heart transplantation in patients seventy years of age and older: A comparative analysis of outcome. J Thorac Cardiovasc Surg. 2001;121:532–41.

10. Dahlberg K, Jaensson M, Eriksson M and Nilsson U. :Evaluation of the Swedish Web-Version of Quality of Recovery (SwQoR): Secondary Step in the Development of a Mobile Phone App to Measure Postoperative Recovery. JMIR research protocols. 2016;5:e192.

11. Jaensson M, Dahlberg K, Eriksson M and Nilsson U. Evaluation of postoperative recovery in day surgery patients using a mobile phone application: a multicentre randomized trial. Br J Anaesth. 2017;119:1030–1038.

12. Halleberg Nyman M, Nilsson U, Dahlberg K and Jaensson M. Association Between Functional Health Literacy and Postoperative Recovery, Health Care Contacts, and Health-Related Quality of Life Among Patients Undergoing Day Surgery: Secondary Analysis of a Randomized Clinical Trial. JAMA Surg. 2018;153:738–745.

13. Myles PS, Weitkamp B, Jones K, Melick J and Hensen S. Validity and reliability of a postoperative quality of recovery score: the QoR-40. Br J Anaesth. 2000;84:11–5.

14. O’Brien SM, Feng L, He X, Xian Y, Jacobs JP, Badhwar V, Kurlansky PA, Furnary AP, Cleveland JC, Lobdell KW, Vassileva C, Wyler von Ballmoos MC, Thourani VH, Rankin JS, Edgerton JR, D’Agostino RS, Desai ND, Edwards FH and Shahian DM. The Society of Thoracic Surgeons 2018 Adult Cardiac Surgery Risk Models: Part 2-Statistical Methods and Results. Ann Thorac Surg. 2018;105:1419–1428.

15. Loughran T and Nagin DS. Finite Sample Effects in Group-Based Trajectory Models. Sociological Methods & Research. 2006;35:250–278.

16. Nagin DS and Odgers CL. Group-based trajectory modeling in clinical research. Annu Rev Clin Psychol. 2010;6:109–38.

17. Savage SA, Sumislawski JJ, Bell TM and Zarzaur BL. Utilizing Group-based Trajectory Modeling to Understand Patterns of Hemorrhage and Resuscitation. Ann Surg. 2016;264:1135–1141.

18. Leffondré K, Abrahamowicz M, Regeasse A, Hawker GA, Badley EM, McCusker J and Belzile E. Statistical measures were proposed for identifying longitudinal patterns of change in quantitative health indicators. J Clin Epidemiol. 2004;57:1049–62.

19. Hartigan JA and Wong MA. Algorithm AS 136: A K-Means Clustering Algorithm. Applied Statistics. 1979;28:100--108.

20. Jones BL, Nagin DS and Roeder K. A SAS Procedure Based on Mixture Models for Estimating Developmental Trajectories. Sociological Methods & Research. 2001;29:374–393.

21. Haviland AM, Jones BL and Nagin DS. Group-based Trajectory Modeling Extended to Account for Nonrandom Participant Attrition. Sociological Methods & Research. 2011;40:367–390.

22. Petersen J, Vettorazzi E, Winter L, Schmied W, Kindermann I and Schäfers HJ. Physical and mental recovery after conventional aortic valve surgery. J Thorac Cardiovasc Surg. 2016;152:1549–1556.e2.

23. Calvert M, Kyte D, Price G, Valderas JM and Hjollund NH. Maximising the impact of patient reported outcome assessment for patients and society. BMJ. 2019;364:k5267.

24. Anderson M, Perrin A. Tech Adoption Climbs Among Older Adults. https://www.pewinternet.org/wp-content/uploads/sites/9/2017/05/PI_2017.05.17_Older-Americans-Tech_FINAL.pdf. Published 2017. Accessed March 30, 2019.

